# Circulating Tumor Cells Are Detectable and Independent of PSA and PSMA-PET Metrics in Localized High-Risk and Biochemically Recurrent Prostate Cancer

**DOI:** 10.1101/2025.07.09.25331014

**Authors:** Shraddha Rastogi, Nahoko Sato, Sunmin Lee, Min-Jung Lee, Yolanda McKinney, Liza Lindenberg, Esther Mena, Anish Thomas, Roshan L. Shrestha, Peter L. Choyke

**Author notes:** Equal contributors (Shraddha Rastogi, Nahoko Sato). Senior corresponding author.

## Abstract

**Purpose:** This study evaluated the feasibility of detecting circulating tumor cells (CTCs) in localized high risk (HR) and biochemically recurrent (BCR) prostate cancer (PCa) patients and examined the correlation between CTCs, serum prostate specific antigen (PSA) levels, 18F-DCFPyL PET parameters and clinical progression.

**Methods:** Baseline samples were collected from 105 PCa patients enrolled in clinical trial NCT03181867 before 18F-DCFPyL PET/CT imaging. Patients were divided into two cohorts-localized HR and BCR. CTCs were enriched using magnetic EpCAM-PE beads and enumerated by flow cytometry (n=82) or by CD45 depletion followed by EpCAM/PSMA expression analysis by ddPCR (n=23). CTC value of ≥3 CTCs/10 mL blood was established as abnormal using 10 healthy female controls. Correlations were assessed between CTCs, PSA, PET parameters and clinical progression.

**Results:** In the flow group, >3 EpCAM⁺ CTCs/10 mL were found in 14% (n=2/14) of HR and 24% (n=16/68) of BCR patients. In the ddPCR group, 53% (n=9/17) of HR and 17% (n=1/6) of BCR patients had greater than normal EPCAM expression, whereas higher than normal PSMA expression was observed in 29% (N=5/17) of HR and 17% (N=1/6) of BCR patients. Furthermore, in the combined cohorts within the ddPCR group, six patients had only EPCAM expression, two had only PSMA and four had both. No significant correlation was found between CTCs and serum PSA or 18F-DCFPyL PET parameters or clinical progression in either group.

**Conclusion:** CTCs were detectable in both HR and BCR patients using flow cytometry and ddPCR based methods even in localized disease, suggesting that CTCs can be used as a liquid biopsy that may reflect underlying disease activity. These findings support the potential role of CTCs as a minimally invasive biomarker independent of serum PSA levels and 18F-DCFPyL PET parameters.

## Introduction

Prostate cancer (PCa) is one of the most prevalent malignancies and ranks as the fifth leading cause of cancer-related deaths among men in the United States [1]. In PCa, localized high-risk (HR) disease represents a subset of patients who have higher risk of disease recurrence based on their PSA and tumor grade [2]. Biochemical recurrence (BCR) in PCa, defined as a rise in serum prostate specific antigen (PSA) levels with no evidence of tumor by conventional imaging, is often observed in patients after initial treatment. Approximately 30% of patients with BCR progress to metastatic disease [3]. Patients in HR and BCR groups have a wide range of outcomes which are influenced by clinical stage, tumor grade, PSA levels, and PSA doubling time. Moreover, prostate specific membrane antigen (PSMA) is significantly overexpressed in most PCa tumors. Recently, PSMA-based positron emission tomography imaging (PSMA-PET) and ^177^Lutetium (Lu)-PSMA-based radioligand therapy in patients with high PSMA expression are emerging as promising tools for managing the disease [4]. Nevertheless, there remains a critical need to identify and validate additional biomarkers to improve treatment decision making and accurately predict disease recurrence in patients.

Liquid biopsy, a rapidly advancing field in oncology, offers great potential to improve cancer diagnosis, monitoring, and treatment selection in a minimally invasive manner that relies on easily accessible patient blood or body fluids. It facilitates the analysis of various biomarkers such as circulating tumor cells (CTCs), circulating extracellular nucleic acids (cell-free DNA; cfDNA), circulating tumor DNA (ctDNA), exosomes, among others [5,6] CTCs are cancer cells which enter the systemic circulation [7]. In most cases, CTCs are extremely rare in blood, with an average 1-10 CTCs per milliliter of blood. However, with more aggressive cancer types, higher cancer stages and other individual patient characteristics higher number of CTCs are detected [8]. Additionally, the phenotypic and molecular heterogeneity of CTCs reflects the underlying tumor’s heterogeneity [9]. Therefore, CTC enumeration and characterization could be effective prognostic markers.

CTCs are commonly identified based on positive epithelial markers such as cytokeratins (Pan-CK) and epithelial cell adhesion molecule (EpCAM), and further defined as negative for the hematopoietic marker, CD45. CTCs can also be characterized based on cancer specific markers. In the context of PCa, CellSearch® is the only Food and Drug Administration (FDA)-approved EpCAM+ CTC detection system for metastatic PCa. The clinical utility of CTCs during therapy for metastatic PCa is recognized [10]. However, the significance of CTCs in localized HR or BCR PCa patients is less clear.

In the present study we have enumerated EpCAM positive CTCs by flow cytometry and analyzed EpCAM and PSMA expression levels on CD45 depleted PBMCs using droplet digital polymerase chain reaction (ddPCR) in treatment naïve localized HR and BCR patients. The aim of this report is to investigate whether CTCs are detectable in these patient cohorts, and to assess if there is a correlation between enriched CTCs, serum PSA levels, clinical progression and 18F-DCFPyL PSMA PET uptake. This exploratory study is a part of a larger clinical imaging study (clinical trial-NCT03181867) that utilizes 18F-DCFPyL, a second generation PSMA-PET agent that binds with high affinity to PSMA.

### Patients and Methodology

#### Patient selection and Study design

Patients ≥18 years with histologically confirmed adenocarcinoma of the prostate were enrolled in clinical trial-NCT03181867 between 04/26/2019 and 08/21/2023 as per international standards of good clinical practice and institutional safety monitoring. Patients met the eligibility criteria for one of the following categories: cohort 1-known localized HR PCa (PSA >10, Gleason 8-10 or clinical stage >T2c) with evidence of disease on conventional imaging or, cohort 2-nonspecific or no evidence of disease on conventional imaging and biochemical PCa relapse defined as PSA > 0.2 ng/ml after surgery or consecutively rising PSA after radiation therapy. In this non-randomized clinical trial, 105 patients underwent a 10 ml blood draw prior to a ^18^F-DCFPyL whole-body PET/CT to investigate the correlation between CTCs and ^18^F-DCFPyL uptake as a part of an exploratory analysis.

Additionally, to set up the threshold value for CTC analysis, blood samples (10 ml) from 10 female healthy donors (HD) ≥ 18 years meeting the eligibility criteria (for donors of human cells, tissues, and cellular and tissue-based product) were also collected anonymously, under protocol number 99-CC-0168 in the Department of Transfusion Medicine, Clinical Center, National Institutes of Health.

CTCs were quantified by two methods: Flow cytometry-involving EpCAM-PE beads enrichment and enumeration and droplet digital PCR (ddPCR), targeting EPCAM and PSMA expression in CTCs enriched by CD45 depletion. Representative workflow for the present study is shown in **Fig. 1**.

**Figure 1:**
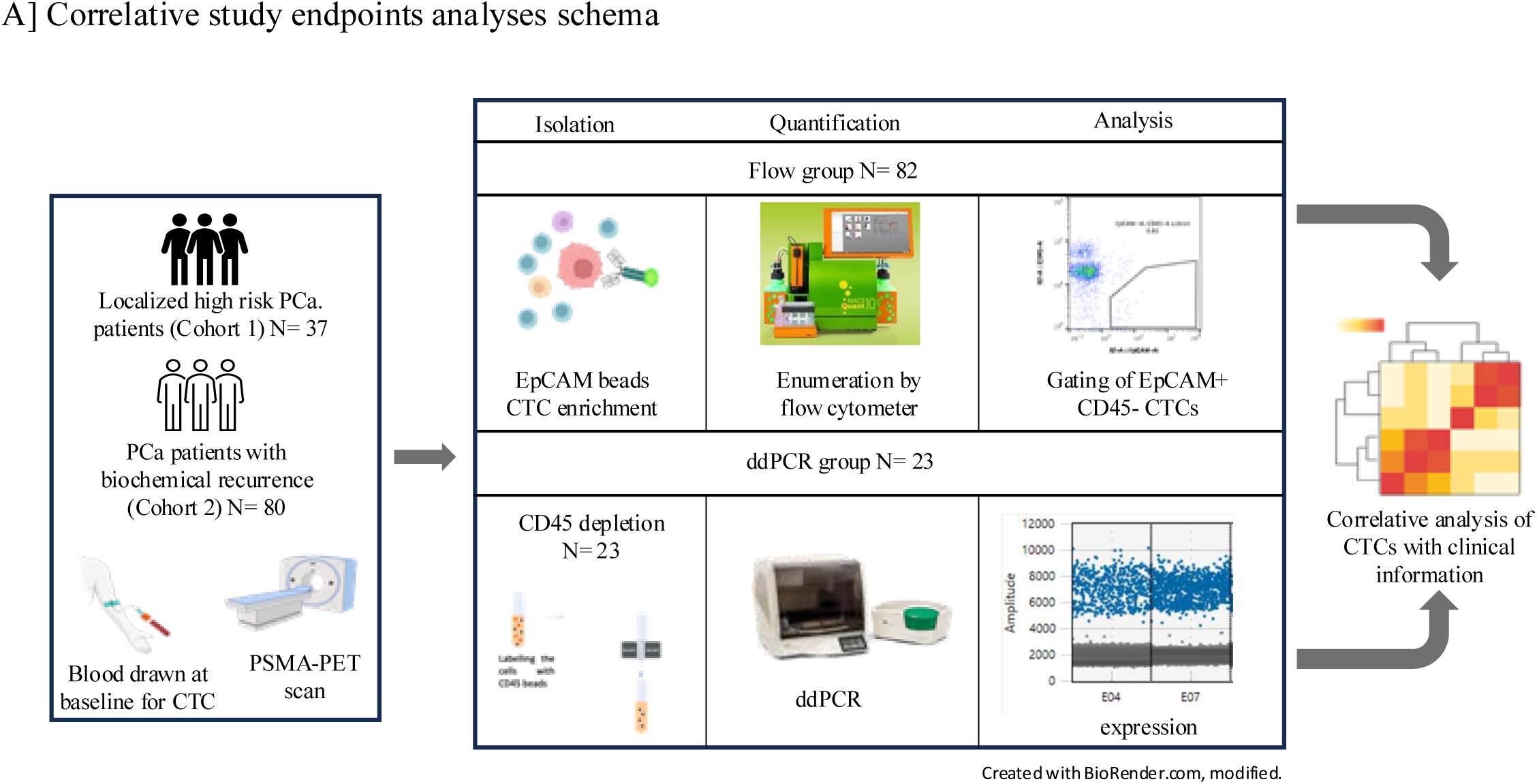
Correlative study endpoint analyses schema: Circulating tumor cells (CTCs) were isolated and quantified from blood samples collected from localized High Risk (HR) and Biochemical Recurrence (BCR) patients. CTC isolation was performed either using EpCAM-based capture or with magnetic CD45 depletion. The quantified CTC data were then systematically correlated with clinical information and PET imaging data to explore associations between CTC burden, imaging findings, and clinical outcomes.

#### CTC isolation by EpCAM beads for flow group

CTC enumeration by flow cytometry was performed in 82 patients-14 from HR and 68 from BCR groups (combined cohorts; flow group) and 10 HD samples. 10ml of peripheral blood was drawn into a CellSave preservative tube (CellSearch, Cat. No. 7900005) for each patient. A detailed procedure for EpCAM positive CTC enumeration by flow cytometry has been published elsewhere [11–13]. Briefly, following RBC lysis, the blood cells were incubated with nuclear dye, Hoechst 33342 (H3570, Life Technologies), followed by the viability dye, LIVE/DEAD Fixable Aqua (L34957, Invitrogen). Cells were then blocked with FcR blocking reagent (130059901, Miltenyi) and incubated with EpCAM microbeads (130-061-101, Miltenyi) on ice for 30 minutes. After incubation, CD45 (clone HI30, BioLegend) and EpCAM (REA764, Miltenyi, Fluorophore-PE) antibodies were added for CTC detection. The EpCAM positive CTCs were detected using magnetic pre-enrichment and multiparameter flow cytometry. Using the gating strategy shown in **Fig. S1A**, viable, nucleated, EpCAM-positive, CD45-negative CTCs were gated and reported as CTC count per 10ml blood. A threshold of CTC> 3 was established from HD samples **(Fig. S1B).**

#### CTC isolation by CD45 depletion for ddPCR group

CTCs were analyzed by ddPCR in 23 patients, (combined cohorts; ddPCR group). Peripheral blood was drawn from each patient into EDTA tubes (BD Vacutainer®; Franklin Lakes, NJ, USA) and was immediately processed for CTC isolation by CD45 depletion. 10 mls of whole blood per patient were centrifuged to remove plasma, followed by ACK RBC lysis. The cells were then washed in cold PBS, incubated on ice for 15 minutes with human Fc-receptor blocking reagent (130059901, Miltenyi Biotec) and microbeads conjugated to monoclonal anti-human CD45 antibody (isotype: mouse IgG2a) (130045801, Miltenyi Biotec). After washing, the cell pellet was resuspended in PBS and loaded onto a MACS® LD Column (130-042-902, Miltenyi Biotec) placed in the magnetic field of a MACS Separator. CD45+ cells were retained within the column, whereas unlabeled cells ran through and were collected as the CD45 depleted cell fraction. After spinning, the cells were stored in an RLT buffer for RNA isolation.

### Total RNA extraction

The total RNA was extracted from the isolated cell pellets stored in RLT buffer using RNeasy Micro Kit (74004, Qiagen) following the user manual instructions without DNase treatment in the automated Qiagen QIAcube connect.

### Droplet Digital PCR (ddPCR)

The total amount of isolated RNA from each patient was converted into cDNA using the Maxima™ H Minus Reverse Transcriptase (RT) Mastermix (M1661, Invitrogen). First, using the TaqMan® PreAmp Master Mix (*4391128, Thermo Fisher Scientific)* a gene-specific pre-amplification for 10 cycles was performed. This was followed by ddPCR on the Bio-Rad QX200 Droplet Digital PCR System with standard thermal cycling parameters (50 °C for 2 min; 95 °C for 10 min followed by 40 cycles of 95 °C for 15 s and 60 °C for 1 min) in a 22 μL total reaction volume using Bio-Rad ddPCR supermix for probes and exon spanning TaqMan™ assays (EpCAM Hs00158980_m; PSMA Hs00379515_m1; Life Technologies). The data was analyzed using QuantaSoft software (Bio-Rad Laboratories, Inc.). The thresholds for the detection of EPCAM and PSMA were set manually (mean+ 2x standard deviation) [14] based on the results from 10 female HD samples **(Fig. S1C).**

### PSMA and EpCAM sensitivity testing using prostate cancer cell line-22Rv1

The sensitivity of ddPCR for PSMA and EPCAM was confirmed by a standard calibration curve of ddPCR concentrations against serially diluted RNA extracted from 22Rv1 cells followed by cDNA synthesis and the pre-amplified cDNA samples. The *R*^2^ values for the correlation between the copies per microliter and initial RNA amount were 0.9909 for PSMA and 0.9999 for EPCAM **(Fig. S1D).**

### Statistical Analysis

Data were analyzed and visualized, and statistical comparisons were performed with R (cran.r-project.org). The Spearman correlation coefficient was calculated for correlation analyses. The heatmaps were generated using the heatmap package. P values <0.05 were considered significant.

## Results

### Patient characteristics of the flow group

The patient characteristics of the flow group are shown in Table 1. Eighty-two patients with the median age of 66.50 years old enrolled for the flow group. At the day of blood draw, clinical and pathological tumor stages were T1: 29.2%, T2: 31.7%, T3: 36.5%, T4:1.6%of the patients. Fourteen of 82 patients (16.86%) were HR (cohort 1) and the remaining 68 (83.13%) were considered BCR (cohort 2). In cohort 1, the tumor stage in majority of patients was T1 (10/14), whereas, in cohort 2, the most common tumor stage was T3 (27/68), followed by T2 (25/68). The median Gleason scores were 8 and 7 in cohort 1 and cohort 2, respectively, with a range of 6-10 in both cohorts. The median serum PSA levels in cohort 1 were 13.01 ng/ml (range of 0.21 to 925.9 ng/ml), while in cohort 2 the median was 1.96 ng/ml (range of 0.2 to 33.21 ng/ml). All patients in cohort 1 were treatment naïve. In cohort 2, 46 of 68 patients had undergone prostatectomy, and 22 patients had undergone radiation therapy.

### CTC detection in flow group

EpCAM+ CD45-CTCs were enumerated in all 82 patients from both cohorts, referred to as the “flow group” **(Fig. 1)**. To determine the threshold for CTC count, EpCAM+ CD45-CTCs were enumerated in 10 normal females (See method section for donor selection). The static cut off for absolute CTC count was set at 3 CTCs/10 ml of whole blood **(Fig. S1B)**. Among 82 patients from both cohorts, 22% (n=18) had more than three EpCAM+ CD45-CTCs/10 ml of whole blood. The remaining 78% (n=65) had CTCs equal to or less than three EpCAM+ CD45-CTCs **(Fig. 2A and 2B).** The range of EpCAM+ CD45-CTCs in the positive flow group was 0-25 CTCs **(Fig. 2B).** When analyzed across individual cohorts, 14% (2/14) and 24% (16/68) of the patients had more than 3 CTCs/10 ml whole blood in the HR and BCR cohorts, respectively **(Fig. 2C),** suggesting that CTCs are more prevalent in the BCR cohort than in the HR cohort.

**Figure 2.**
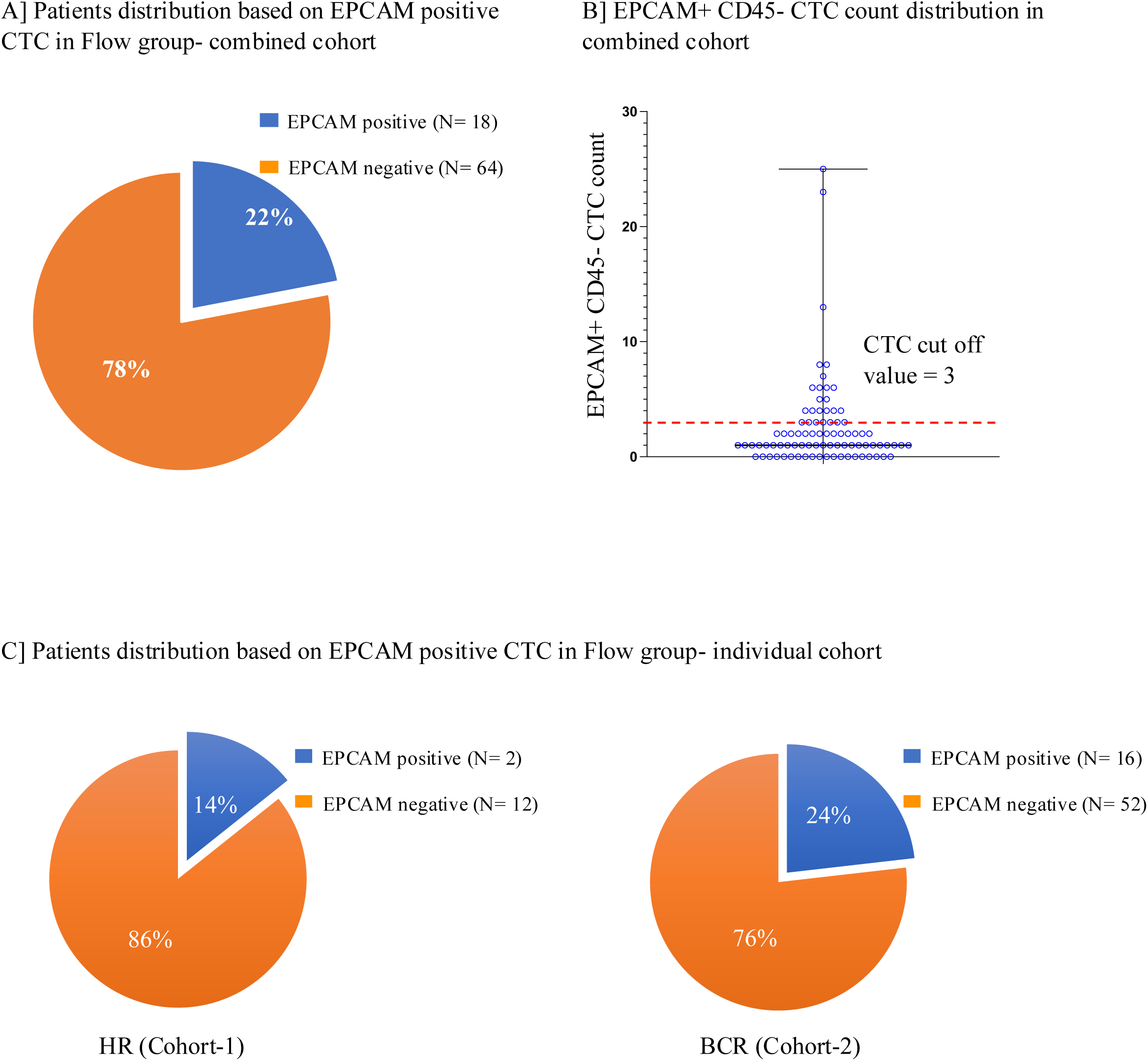
Detection of EpCAM-positive CTCs in the flow group. **(A)** Proportion of patients with detectable EpCAM-positive CTCs across the combined cohorts. **(B)** Distribution of EpCAM-positive CTC counts in individual patients, the maximum observed count was 25 CTCs, and a positivity threshold of ≥3 CTCs was applied based on HD EpCAM positivity, as defined in Supplementary Figure S1. **(C)** Frequency of EpCAM-positive CTCs in HR group (Cohort 1) (left panel) and BCR group (Cohort 2) (right panel).

### Patient characteristics of the ddPCR group

The patient characteristics of the ddPCR group (from both cohorts) are shown in Table 2. Twenty-three patients with a median age of 67 years were enrolled in the ddPCR group and divided into two cohorts, 17 of 23 (74%) were from the HR cohort (cohort 1), while 6 (26%) were from the BCR (cohort 2). On the day of blood draw, clinical and pathological tumor stages were T1: 21.7%, T2: 34.78% and T3: 43.47% in this group. In cohort 1, the tumor stage in the majority of patients was T3 (9/17), whereas, in cohort 2 (BCR), the predominant tumor stage was T2 (4/6). The median Gleason scores were 7 in cohort 1 and 8 in cohort 2, respectively, with a range of 6-9 in both cohorts. The median serum PSA level in cohort 1 was 14.0 ng/ml (range: 5.6-37.4 ng/ml), whereas in cohort 2, the median serum PSA level was 0.45 ng/ml (range: of 0.3-4.8 ng/ml). Although the absolute range of PSA levels is different, the pattern of PSA values is similar between the flow group and the ddPCR group (Table 1 and Table 2). All patients in cohort 1 (N=17) were treatment-naïve, whereas all patients in cohort 2 had undergone either prostatectomy (5/6) or radiation therapy (1/6) (Table 2).

### EPCAM expression by ddPCR in isolated CTCs

Next, we utilized ddPCR to examine the EPCAM expression in CTCs enriched post CD45 depletion in 23 patients from both cohorts, referred to as the “ddPCR group.” CD45 depleted cells from whole blood were used for this analysis **(Fig. 1)**. To determine the threshold cut-off value for EPCAM expression, ddPCR analysis was performed in the 10 female HD cohort (See method section for donor selection). Based on this analysis, 30.52 copies/μl was set as the cut-off value for EPCAM expression **(Fig. S1C)**. Among 23 patients from the combined cohorts, 43% (N=10) had EPCAM expression levels higher than the cut-off value with a range of 35.7-75.13 copies/ μl **(Fig. 3A and 3B).** When analyzed individually, 53% (9/17) of patients in the HR cohort and 17% (1/6) in the BCR cohort had positive EPCAM expression **(Fig. 3B and 3C)**. The median EPCAM expression level in CD45 depleted cells was three times higher in patients from the HR cohort than the BCR cohort **(Fig. 3B).**

**Figure 3.**
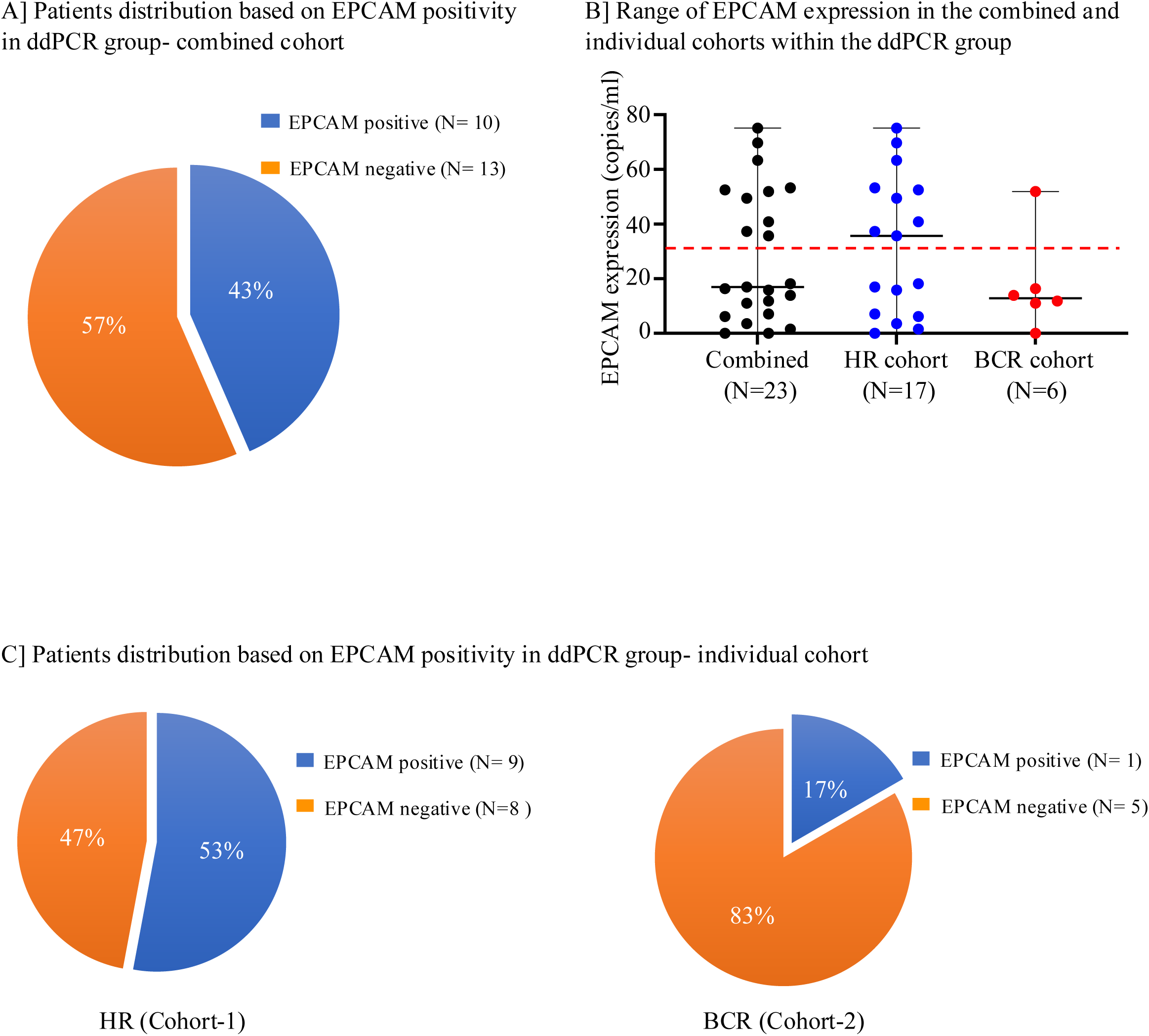
Detection of EpCAM expression by ddPCR in CD45-depleted cells from patient cohorts. **(A)** Proportion of patients with detectable EpCAM expression assessed by ddPCR across the combined cohorts. **(B)** Distribution of EpCAM expression levels (copies/µL) in individual patients, shown for both combined and individual cohorts. The red line indicates the threshold for EpCAM positivity. **(C)** Frequency of EpCAM-positive cases in HR group (Cohort 1) (left panel) and BCR group (Cohort 2) (right panel).

### PSMA expression by ddPCR in isolated CTCs

PSMA is a biomarker for PCa and is the binding target of ^18^F DCFPyl PET scans, therefore, the expression levels of PSMA were examined in CTCs isolated from both cohorts in the ddPCR group. The threshold cut-off value was determined using the same HD cohort as the EpCAM expression analysis. Based on this analysis 8.05 copies/μl was set as the threshold cut-off value **(Fig. S1C)**. Among 23 patients from the combined cohorts, 26% (N=6) had PSMA expression levels higher than the cut-off value, with a range of 10.5-37.85 copies/ μl **(Fig. 4A and 4B)**. When analyzed individually, 29% (5/17) and 17% (1/6) of the patients had positive PSMA expression in the HR and the BCR cohorts, respectively **(Fig. 4B and 4C).** Next, we examined whether there is a correlation between EPCAM and PSMA expression in patients from the combined cohorts in the ddPCR group. Among the 16 patients who had either EPCAM or PSMA expression above the cut-off, only 25% (N=4) had high expression of both EPCAM and PSMA **(Fig. 4D)**, suggesting that EPCAM and PSMA do not necessarily co-express in CTCs from HR and BCR PCa patients.

**Figure 4.**
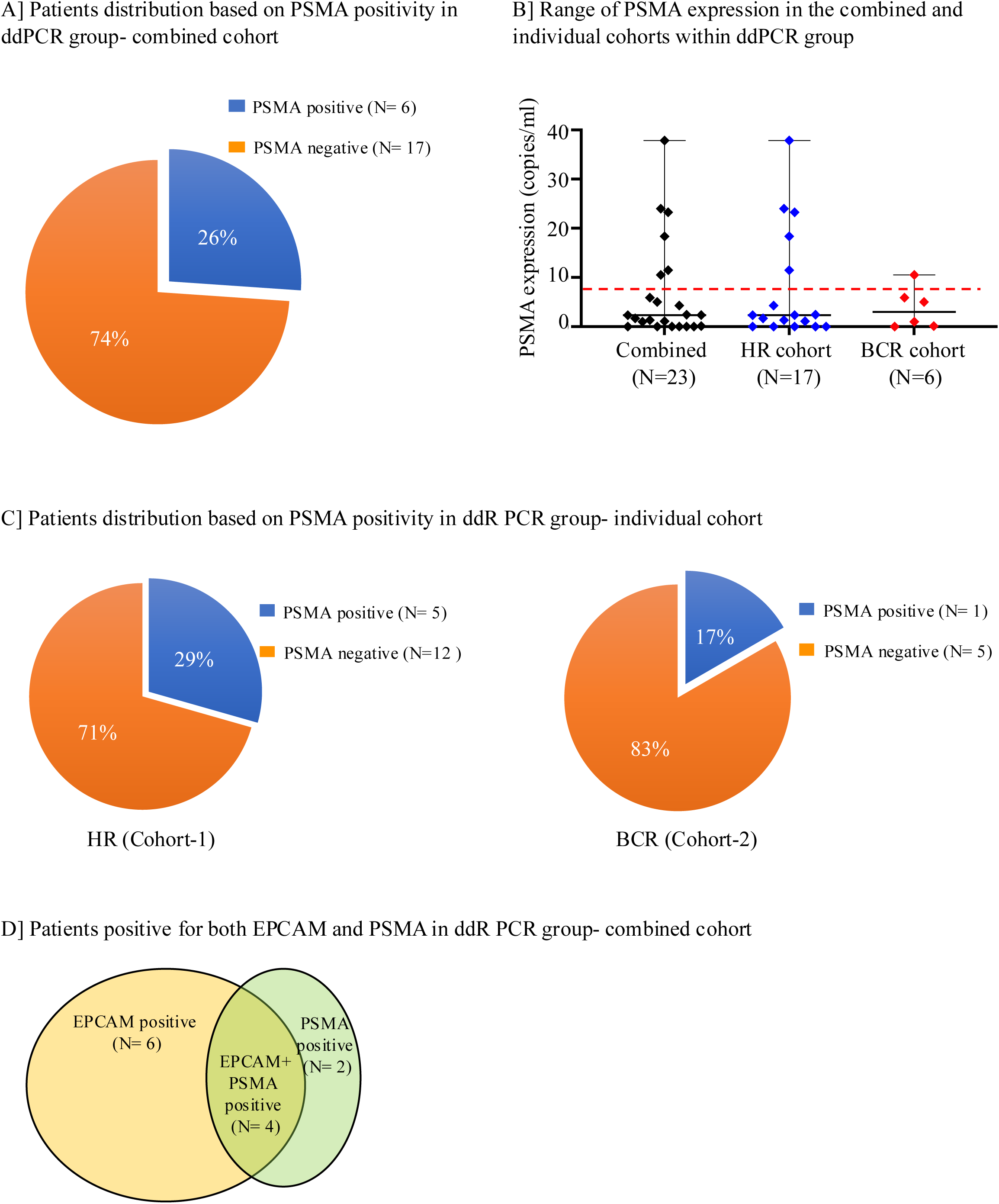
Detection of PSMA expression by ddPCR in CD45-depleted cells from patient cohorts. **(A)** Proportion of patients with detectable PSMA expression assessed by ddPCR across the combined cohorts. **(B)** Distribution of PSMA expression levels (copies/µL) in individual patients, shown for both combined and individual cohorts. The red line indicates the threshold for PSMA positivity. **(C)** Frequency of PSMA-positive cases in HR group (Cohort 1) (left panel) and BCR group (Cohort 2) (right panel). **(D)** Venn diagram illustrating the distribution of EpCAM-positive, PSMA-positive, and dual EpCAM+PSMA+ patients within the ddPCR cohort.

### Correlative analysis of CTCs with clinical progression in combined cohort

We analyzed the correlation of clinical progression and CTC count or EPCAM/PSMA expression. The mean and range of follow up for clinical progression monitoring was 33.5 months (22 days – 70.7 months) for flow group and 20.1 months (13.1 months – 26.4 months) for ddPCR group. In the flow group, 31 patients did not progress, 31 patients had clinical signs of progression, and 20 patients were not monitored for progression. To evaluate if CTC count could be used as a predictor for progression, the number of EpCAM+ CD45-CTCs were analyzed against clinical outcome (progression or no progression). CTC count more than cut off i.e. 3 CTCs/10ml was observed in 22.5 % (7/31) of patients with progression, compared to that in 16.1% (5/31) of patients with no progression **(Fig. 5)**. Overall, there was no significant difference in the number of CTCs in patients with or without progression.

**Figure 5.**
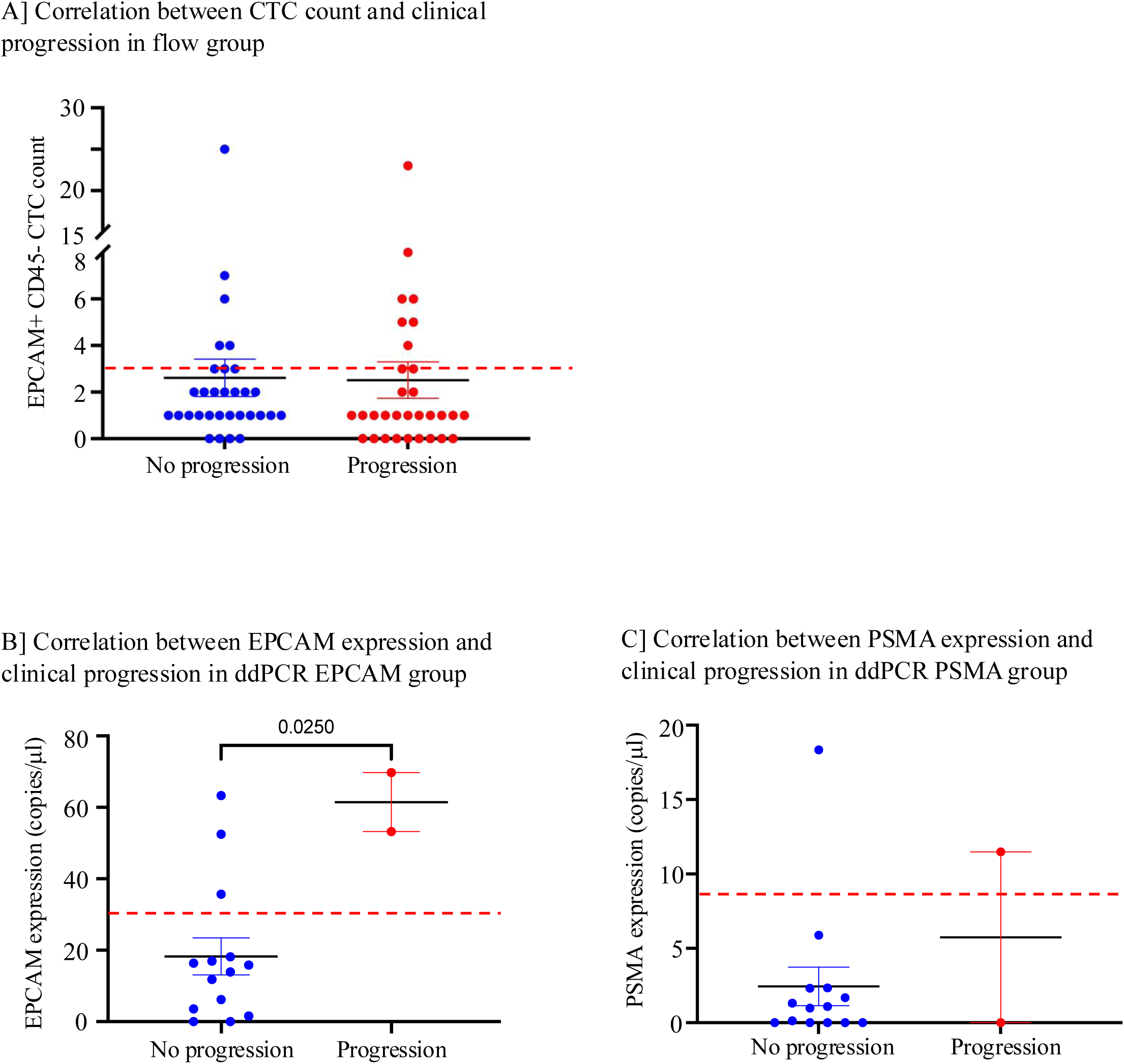
Correlation between CTC measures and clinical progression in Flow group and ddPCR group. (**A**) Comparison of EpCAM-positive CTC counts between patients with no clinical progression and those with progression in the flow group. The dotted line indicates the CTC positivity threshold (≥3). **(B)** Comparison of expression levels of EpCAM (measured by ddPCR) in CD45-depleted cells, between patients with no clinical progression and those with progression in the ddPCR group. **(C)** Comparison of expression levels of PSMA (measured by ddPCR) in CD45-depleted cells between patients with no clinical progression and those with progression in the ddPCR group

In the ddPCR group, 14 patients were stable, 2 patients progressed and the remaining 7 were not monitored for progression. Two patients that progressed had higher EPCAM expression than the cut off value of 30.2 copies/ml **(Fig. 5B)**. A majority of patients who did not progress (11/14) had EPCAM expression levels lower than the cut off. Only one of 14 patients without progression had higher PSMA expression than the cut off value of 8.0 copies/ml and one of two patients with progression had higher PSMA expression **(Fig. 5C)**. In this group statistical evaluation was limited due to uneven distribution of patients in progressed versus not progressed categories.

Next, we evaluated if CTC count correlated with the clinical and pathological tumor stages. We did not observe a correlation either in the flow group or ddPCR group, suggesting that CTC count is not a predictive marker for tumor staging in HR and BCR prostate cancer patients.

### Correlative analysis of CTCs with PSA and ^18^F-DCFPyL uptake in combined cohorts

To understand the correlation between EpCAM+ CTC count in the flow group **(Fig. 6A and 6B)** as well as the expression of EPCAM and PSMA in the ddPCR group **(Fig. 6C and 6D)** with PSA levels and ^18^F-DCFPyL uptake, a heatmap and a correlogram was generated for each group. We did not observe correlations between PSA levels, SUV total, total PSMA lesion volume, total Prostate (sum of all prostate/bed_volumes), SUVmax, and total volume, and either CTC count or expression of EPCAM/PSMA **(Fig 6B and 6D)**.

**Figure 6.**
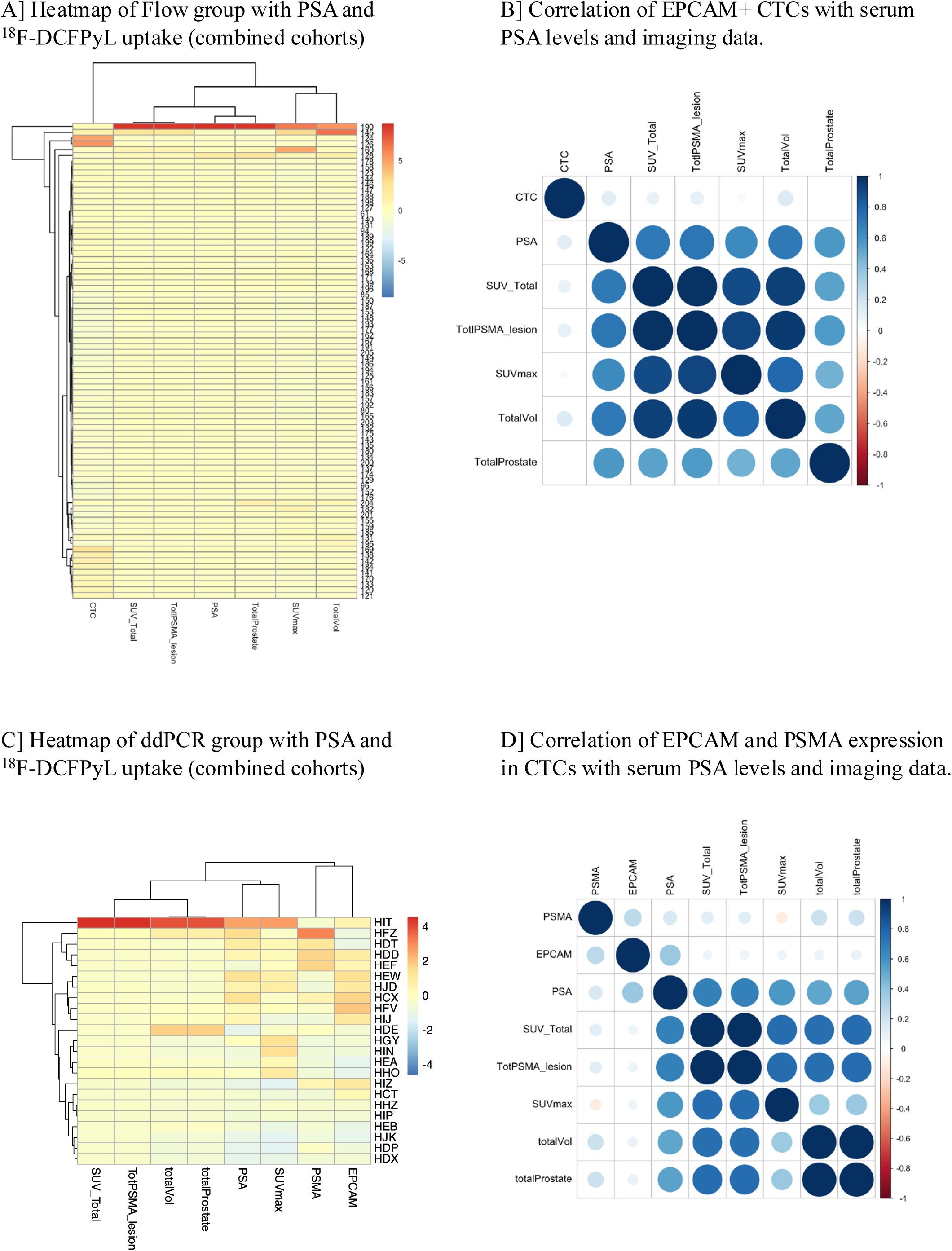
Integration of CTC biomarkers and PET imaging parameters in Flow group and ddPCR group. **(A)** Heatmap of the flow cytometry group showing PSA levels and 18F-DCFPyL PET imaging parameters. The longitudinal axis represents individual patients, while the transverse axis includes CTC count, PSA levels, and PET imaging metrics such as SUVtotal, total PSMA lesion burden, total prostate volume, SUVmax, and total lesion volume. Each cell is color-coded based on the magnitude of measured values within the given range. **(B)** Correlogram illustrating correlations between EpCAM positive CTSs (Flow group) and PET imaging/biomarker parameters. Positive correlations are shown in blue, negative in red. Circle size and color intensity correspond to the correlation coefficient magnitude. A color-coded reference scale is shown on the right. **(C)** Heatmap of the ddPCR group displaying EpCAM expression, PSMA expression and imaging parameters as in panel A. CD45-depleted RNA was used for EpCAM and PSMA quantification. Data are shown across combined patient cohorts. **(D)** Correlogram representing the correlations between EpCAM expression (ddPCR group) and PET imaging/PSA parameters. Visual representation follows the same scheme as in panel B.

## Discussion

Several studies have reported CTCs in patients with localized prostate cancer using different isolation methods. However, to the best of our knowledge, this study represents one of the first attempts to elucidate a correlation between CTCs and PSMA PET i.e ^18^F-DCFPyL uptake in localized HR and BCR prostate cancer patients. In localized HR / BCR prostate cancer patients there are relatively few CTCs therefore, one of the exploratory objectives of this clinical trial was to investigate whether CTC count can be used as a clinical marker in lieu of repeated PSMA PET scans. Dynamic monitoring of CTCs has been widely used in cancer clinical trials for prognosis and patient stratification [7], [15] however, due to their rarity, isolation and detection of CTCs in early stage prostate cancer is quite challenging.

The collective range of CTCs in this study was 0 to 25. Using the same enrichment method, we detected CTCs ranging from 0 to 2107/10ml blood (N=17) in our previous phase 2 mCRPC study [11]. In the ddPCR group, EPCAM expression was 47% in HR and 17% in BCR. Regardless of the isolation technique, the percentage of EpCAM-positive CTCs in HR was in line with the other studies where the positive rate of EpCAM+ CTC was reported as 11-21% in HR PCa patients. Furthermore, like the present study, these studies did not demonstrate an association of CTCs with either prognostic factors or clinicopathological characteristics [16–20].

PSMA PET imaging has become an important biomarker in PCa. Recently, PSMA-PET has become a central diagnostic tool in staging and treatment decision. ^177^Lu-PSMA-617, a PSMA directed radioligand treatment for metastatic castration-resistant prostate cancer has been approved by the US-FDA for clinical practice [21]. Therefore, examining PSMA expression in CTCs may identify a potential biomarker for PSMA-targeted therapeutics. In this study, 29% of the patients in HR and 17% in BCR had PSMA expression above the normal cutoff by ddPCR. A previous study reported that 50-70% of CTCs were positive for PSMA expression in localized PCa patients [22]. The same study further checked the BCR free survival in these patients’ post-surgery and observed a trend toward higher PSMA expression in BCR positive patients than in BCR-negative patients.

In the ddPCR group, co-expression analysis of PSMA and EPCAM in CD45 depleted cells showed that some patients had only EPCAM expression, others had only PSMA expression, and some patients showed co-expression of PSMA and EPCAM. These results suggest that in addition to serving as a prognostic tool, gene expression analysis on CTCs also may provide information on tumor heterogeneity. Heterogeneity in PSMA expression levels within tumors has also been previously reported [23–25].

Higher CTC count is linked with the estimate of disease progression for several cancer types including breast, mCRPC and colorectal but typically in the metastatic setting when more CTCs are present [26–28]. In this study, we also correlated clinical progression with EpCAM+ CD45-CTC counts or high EPCAM/PSMA expression. The number of patients with and without progression in the flow group was comparable, 31 patients in each arm, whereas, in the ddPCR group, the number of patients with progression was only two vs. 14 without progression within the monitoring time range of 13.1 months – 26.4 months. We did not observe any significant correlation between CTC count and clinical correlation in both cohorts possibly due to certain limitations discussed below.

There are several limitations to this study that prevent a conclusion on whether CTC count and EPCAM/PSMA expression could be used as the predictive biomarker for clinical outcome. First, the fate of disease outcome in non-progressed patients that had higher CTC count or EPCAM/PSMA expression is not known and takes many years to ascertain in the case of prostate cancer. Second, the number of patients with progression in the ddPCR group was too low to compare with patients without progression. Hence, a greater number of progressed patients and prolonged disease monitoring is warranted in future studies. It is also important to note that this study is limited by the small number of patients in the ddPCR group. No direct comparison was made between the two techniques, as the enrichment and detection strategies were different. Furthermore, as patients were accrued from an imaging trial, blood samples for CTCs were only collected before the PSMA-PET scan and the patients were not followed up. EpCAM as a surface marker also has limitations in detecting CTCs, particularly those undergoing epithelial-to-mesenchymal transition (EMT). During EMT, CTCs can downregulate their expression of EpCAM, potentially evading detection by EpCAM-based methods [29,30]. The sensitivity of the CTC detection rate depends on the method used to isolate and detect CTCs. Recently, Cho et al. reported a 75.5% EpCAM+ CTC detection rate in localized PCa patients, using a microfluidic tool, CTC-dμChip [22]. Andra Kuske et. al. reported improved detection of CTCs in non-metastatic HR PCa. They found that CTCs were detected in 37%, 54.9%, and 58.7% of patients using CellSearch, CellCollector, and EPISPOT, respectively. Additionally, they observed a significant difference in CTC capture before and after surgery in patients using CellCollector [31]. This suggests that integration of different markers for CTCs and sensitive CTC isolation techniques is pivotal for CTC detection and analysis in localized PCa. CTCs with both epithelial and mesenchymal characteristics have been identified in patients with PCa using the cell size-based Parsortix isolation system [32]. Another approach to detect CTCs was developed based on telomerase-specific, replication-selective oncolytic herpes-simplex-virus-1 that targeted telomerase-reverse-transcriptase-positive cancer cells and expressed green-fluorescent-protein that identified viable CTCs. Identification of CTCs using this approach was tested in lung, colon, liver, gastric, pancreatic cancer, and glioma where CTC-positive rates increased with tumor progression [33].

In the present study, EpCAM+ CTCs or EPCAM and PSMA expression levels were not associated with either PSA level or ^18^F-DCFPyL uptake. Lack of association between CTC and PSA levels was consistent with a study by Meyer et al. that reported no correlation between CTCs isolated using Cell Search system and PSA in early BCR patients [19]. In contrast, CTCs detected by EPISPOT in non-metastatic HR PCa patients before surgery significantly correlated to PSA levels and clinical tumor stage [31]. The discrepancies in CTC detection and correlation across different studies could be dependent on the surface antigen used to isolate CTCs. For example, CellSearch uses EPCAM as a surface marker, whereas EPISPOT is an EPCAM-independent assay that detects PCa CTCs based on active secretion of PSA.

In conclusion, this study highlights the potential of CTC detection in localized HR and BCR PCa patients. The presence of either or both EpCAM+ and PSMA+ CTCs underscores the underlying degree of tumor heterogeneity in prostate cancer and the need for multiple markers. While no significant correlations were found between CTCs and PSA levels or 18F-DCFPyL PET parameters, the study emphasizes the importance of sensitive isolation techniques and the integration of various markers for improved CTC detection. Further research with larger cohorts and long-term follow-up is needed to establish the prognostic significance of CTCs in these patient populations.

## Figure Legends

**Supplementary Figure 1. Threshold determination, gating strategy, and assay validation for EpCAM and PSMA detection**

**(A)** Representative gating strategy used for the detection of EpCAM-positive circulating tumor cells (CTCs) by flow cytometry. From the nucleated viable cells, CD45-negative events were gated and further analyzed for EpCAM positivity. **(B)** Summary table establishing a CTC count threshold of ≥3 for positivity in the flow group. The cutoff was determined based on analysis of female healthy donors (HD), who served as negative controls. **(C)** Threshold determination for EpCAM and PSMA expression levels in the ddPCR group. CD45-depleted RNA from female healthy donors was used to establish background expression, and thresholds were calculated as mean + 2 standard deviations (2SD). **(D)** Sensitivity testing for EpCAM and PSMA detection using 22Rv1 prostate cancer cells as a positive control. This assay confirms the ability of ddPCR platform to reliably detect marker expression at known positive levels.

**Table 1. Baseline clinical and demographic characteristics of patients in the Flow group.**

This table summarizes key clinical and demographic features of patients in the Flow group, including age, gender, disease status, and relevant laboratory parameters such as Gleason score and PSA levels at study entry.

**Table 2. Baseline clinical and demographic characteristics of patients in the ddPCR group.**

This table presents the key clinical and demographic characteristics of patients in the ddPCR group, including age, gender, disease status, and relevant laboratory parameters such as Gleason score and PSA levels at the time of study entry.

## Data Availability

All data produced in the present study are available upon reasonable request to the authors

## Acknowledgement

We gratefully acknowledge Jane Trepel, whose initial idea and collaborative insights were instrumental in conceptualizing this research. We sincerely thank the patients and healthy donors whose contributions were crucial for conducting this research.

This study was supported by the Center for Cancer Research, the Intramural Program of the NCI (ZIA BC 010655 and ZIC BC 012174). The content of this publication does not necessarily reflect the views or policies of the Department of Health and Human Services, nor does mention of trade names, commercial products, or organizations imply endorsement by the U.S. Government.

## Author Contributions

Conceptualization, P.L.C.; Sample handling, S.R, N.S.; Data collection, analysis and interpretation, S.R, N.S, S.L., M.J.L., L.L., E.M.; writing-original draft, S.R., N.S.; writing-review and editing, S.L., M.J.L.,A.T.,R.L.S.,P.L.C.

