## Supplemental Figure 1 for "Circulating Tumor Cells Are Detectable and Independent of PSA and PSMA-PET Metrics in Localized High-Risk and Biochemically Recurrent Prostate Cancer"

### Figure S1

#### A] Gating strategy of Flow group for EPCAM positive CTCs

Nucleated cell gating → Gated out non-viable cells → Gated out small cells → Gated out hematopoietic cells

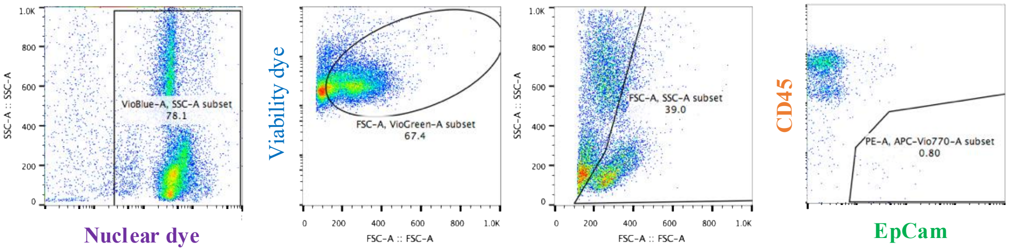


#### B) Flow group

| Healthy Donor | EpCam+ Cell Count |
|---|---|
| HD1F | 3 |
| HD2F | 3 |
| HD3F | 0 |
| HD4F | 0 |
| HD5F | 1 |
| HD6F | 0 |
| HD7F | 0 |
| HD8F | 1 |
| HD9F | 0 |
| HD10F | 0 |

#### C] ddPCR group

| Samples | Target | Conc. (copies/µL) | Target | Conc. (copies/µL) |
|---|---|---|---|---|
| 22Rv1 | EPCAM | 6,558.11 | PSMA | 4,870.48 |
| HD1D | EPCAM | 0.03 | PSMA | 0.00 |
| HD2D | EPCAM | 2.67 | PSMA | 0.00 |
| HD3D | EPCAM | 7.03 | PSMA | 0.00 |
| HD4D | EPCAM | 4.10 | PSMA | 0.00 |
| HD5D | EPCAM | 2.74 | PSMA | 0.00 |
| HD6D | EPCAM | 19.52 | PSMA | 0.00 |
| HD7D | EPCAM | 0.03 | PSMA | 8.59 |
| HD8D | EPCAM | 0.00 | PSMA | 1.75 |
| HD9D | EPCAM | 28.07 | PSMA | 3.94 |
| HD10D | EPCAM | 24.28 | PSMA | 5.42 |
| NTC | EPCAM | 0.09 | PSMA | 0.00 |

| | EpCAM | PSMA |
|---|---|---|
| Mean | 8.85 | 1.97 |
| Standard deviation | 10.84 | 3.04 |
| Threshold (Mean + 2*Standard dev) | 30.52 | 8.05 |

#### D] EpCAM and PSMA sensitivity testing using 22Rv1 cell line

| RNA (pg) | EPCAM (copies/ul) |
|---|---|
| 1000 | 3,505.86 |
| 200 | 702.67 |
| 40 | 173.06 |
| 8 | 22.41 |
| 4 | 10.14 |
| 2 | 6.38 |
| 1 | 2.45 |
| 0 | 0.00 |

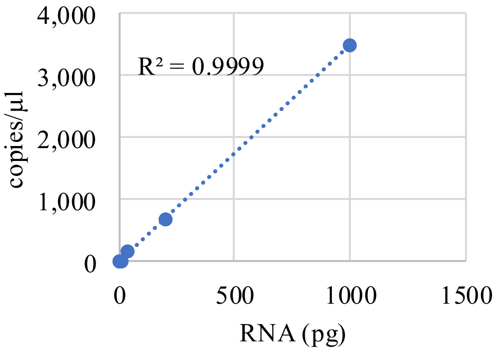


| Without amplification | | Copies/µl |
|---|---|---|
| Rv1_100ng | EPCAM | 340.63 |
| Rv1_100ng | PSMA | 1,899.24 |

| RNA (pg) | PSMA (copies/ul) |
|---|---|
| 1000 | 12,478.73 |
| 200 | 3,670.80 |
| 40 | 860.58 |
| 8 | 121.35 |
| 4 | 55.74 |
| 0 | 0.00 |

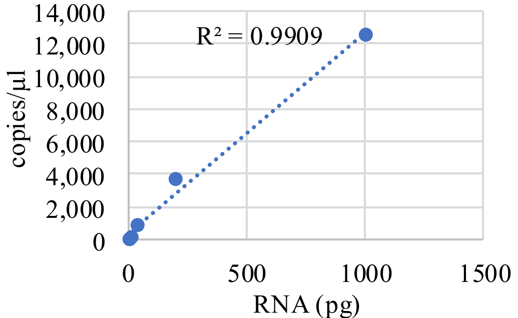
