## Supplementary material for "Circulating Tumor Cells Are Detectable and Independent of PSA and PSMA-PET Metrics in Localized High-Risk and Biochemically Recurrent Prostate Cancer": Tables

Table 1: Patient characteristics- Flow group

| **Characteristics** | | |
|---|---|---|
| Median age of combined cohorts (years old) | | 66.50 [48-85] [min-max] |
| High Risk (Cohort 1) | | N= 14 (16.86%) |
| Biochemical recurrence (Cohort 2) | | N= 68 (83.13%) |
| Clinical or pathological T stage at the day of blood drawn | | T1c: N= 24 (Cohort 1 N= 10, Cohort 2 N= 14)<br>T2: N= 26 (Cohort 1 N= 1, Cohort 2 N= 25)<br>T3: N= 30 (Cohort 1 N= 3, Cohort 2 N= 27)<br>T4: N= 2 (Cohort 1 N= 0, Cohort 2 N= 2) |
| Median original Gleason score | Cohort 1 | 8 [6-10] [min-max] |
| | Cohort 2 | 7 [6-10] [min-max] |
| Median PSA level (ng/ml) | Cohort 1 | 13.01 [0.21- 925.9] [min-max] |
| | Cohort 2 | 1.96 [0.2-33.31] [min-max] |
| No treatment (Cohort 1) | | N= 14 |
| Pre-treatment (Cohort 2) | | Prostatectomy N= 46<br>Radiation therapy N= 22 |
| Median EpCAM CTC count | Cohort 1 | 2 [0-13] [min-max] |
| | Cohort 2 | 1 [0-25] [min-max] |
| Progression since date drawn to last available visit | Cohort 1 | Yes (N= 4), No (N= 6), N/A (N= 4) |
| | Cohort 2 | Yes (N= 27), No (N= 25), N/A (N= 16) |

Table 2: Patient characteristics- ddPCR group

| Characteristics | | |
|---|---|---|
| Median age of combined cohorts (years old) | | 67 [51-81] [min-max] |
| High Risk (Cohort 1) | | N= 17 (73.91%) |
| Biochemical recurrence (Cohort 2) | | N= 6 (26.08%) |
| Clinical or pathological T stage at the day of blood drawn | | T1c: N= 5 (Cohort 1 N= 4, Cohort 2 N= 1)<br>T2: N= 8 (Cohort 1 N= 4, Cohort 2 N= 4)<br>T3: N= 10 (Cohort 1 N= 9, Cohort 2 N= 1)<br>T4: N= 0 (Cohort 1 N= 0, Cohort 2 N= 0) |
| Median original Gleason score | Cohort 1 | 7 [6-9] [min-max] |
| | Cohort 2 | 8 [6-9] [min-max] |
| Median PSA level (ng/ml) | Cohort 1 | 14.0 [5.6-37.4] [min-max] |
| | Cohort 2 | 0.45 [0.3-4.8] [min-max] |
| No treatment (Cohort 1) | | N= 17 |
| Pre-treatment (Cohort 2) | | Prostatectomy N= 5<br>Radiation therapy N= 1 |
| Median EPCAM and PSMA expression (copies/µl ) | Cohort 1 | EPCAM: 35.7 [0-75.13] [min-max]<br>PSMA : 2.32 [0-37.85] [min-max] |
| | Cohort 2 | EPCAM: 12.86 [0-51.93] [min-max]<br>PSMA: 2.995 [0-10.5] [min-max] |
| Progression since date drawn to last available visit | Cohort 1 | Yes (N= 2), No (N= 10), N/A (N= 5) |
| | Cohort 2 | Yes (N= 0), No (N= 4), N/A (N= 2) |
